# Predicting Car Accident Severity in Northwest Ethiopia: A Machine Learning Approach Leveraging Driver, Environmental, and Road Conditions

**DOI:** 10.1101/2025.01.12.25320441

**Authors:** Abraham Keffale Mengistu, Andualem Enyew Gedefaw, Nebebe Demis Baykemagn, Agmasie Damtew Walle, Tirualem Zeleke Yehuala, Meron Asmamaw Alemayehu, Mengistu Abebe Messelu, Bayou Tilahun Assaye

## Abstract

**Background:** Car accidents in Northwest Ethiopia have significantly increased in severity, with increasing impacts on public safety. This study aims to predict car accident severity in the region by considering driver behavior, environmental conditions, and road characteristics, utilizing machine learning models to enhance traffic safety measures.

**Methods:** This study used a dataset comprising accident records, weather conditions, road types, traffic volume, and driver characteristics. Various machine learning models, including logistic regression, decision trees, random forests, gradient boosting, XGBoost, LightGBM, support vector machines, K-nearest neighbors, multilayer perceptron (neural networks), and naive Bayes, have been employed to predict accident severity. Model performance was assessed in terms of accuracy, precision, recall, and the F1 score.

**Results:** Driver-related factors, including age and behavior, were found to significantly influence accident severity. Environmental factors, such as weather and road type, also play crucial roles in determining outcomes. Among the evaluated models, the random forest classifier demonstrated superior performance, achieving an initial accuracy of 78% in predicting fatal accidents. Its performance was improved to 82% through hyperparameter tuning, highlighting its strong predictive ability.

**Conclusions:** This research highlights the importance of environmental and driver-related factors in predicting accident severity in Northwest Ethiopia. The random forest model proves to be an effective tool for forecasting accident severity, which could inform policies and interventions to improve road safety in the region. Future work should focus on expanding the dataset and exploring additional models for better prediction accuracy.

## Background

Traffic accidents remain a major public health and safety concern worldwide, with varying severities depending on numerous factors, such as driver behavior, road conditions, and environmental influences(1,2). In Ethiopia, road accidents have become an increasingly critical issue, particularly in the northwestern region, where road infrastructure and safety standards are developing(3,4). Despite the growing number of road accidents, limited research has been conducted on predicting accident severity in this context, especially using advanced data-driven ML approaches.

The literature has focused primarily on accident prediction in developed countries, where data availability and road safety measures are more comprehensive(5,6). Studies often highlight the influence of driver attributes, weather conditions, and road infrastructure on accident outcomes, employing statistical methods or basic machine learning models(5–7). However, there is a notable gap in the application of such models to regions such as Northwest Ethiopia, where road safety data are sparse and where environmental conditions differ from those in other regions(4). This necessitates a more localized approach that uses machine learning algorithms to predict car accident severity on the basis of relevant variables such as driver, environmental, and road conditions.

This study aims to fill this gap in machine learning models tailored to Northwest Ethiopia, leveraging data on driver characteristics, environmental conditions, and road infrastructure to predict accident severity. Through this, the study aims to contribute to road safety measures, enabling the identification of key factors influencing accident severity and providing actionable insights to policymakers and traffic authorities. These findings are essential for enhancing preventive strategies, improving traffic regulations, and ultimately reducing the occurrence of severe accidents in the region.

## Methods

### Aim and study design

This study aims to predict the severity of car accidents in Northwest Ethiopia via machine learning techniques and considers various factors, such as driver characteristics, environmental conditions, and road infrastructure. The study employs a retrospective observational design, using data collected from traffic accident reports, weather stations, and road condition databases from the East Gojjam zonal police department and Debre Markos city police office. The research was conducted in Northwest Ethiopia, focusing on regions with high traffic volumes and varying road types, to ensure a representative sample of the diverse conditions affecting road safety.

### Participants and materials

The data for this study were obtained from local traffic accident records, including police reports, which contain detailed information about the time, location, weather conditions, road types, and severity of the accidents. Road conditions, such as road type and traffic volume data, were gathered from local traffic authorities.

### Data collection and preprocessing

The dataset includes an accident recording period year (2018--2023) from northwestern Ethiopia, with over 2000 recorded accidents from the Zonal office and city administration offices. The variables considered in the analysis include driver age, gender, behavior, fatigue, distractions, alcohol influence, seatbelt usage, weather conditions, road conditions, lighting conditions, traffic volume, time of day, and vehicle type. All the data were preprocessed to handle missing values, outliers, and normalization of numerical features. The outcome variable is generated from the column fatalities, which are called fatal with numbers greater than zero and nonfatal rows with zero.

### Data Balancing

In many machine learning applications, datasets can suffer from class imbalance, where one class has significantly fewer instances than others. This imbalance can lead to biased models that favor the majority class, potentially overlooking crucial information in the minority class. To address this issue, various data balancing techniques have been developed, including SMOTE (8–10).

SMOTE is a popular oversampling technique that addresses class imbalance by generating synthetic samples for the minority class. It works by selecting minority class samples and creating synthetic samples along the line segments connecting them to their nearest neighbors in the feature space. This process increases the number of minority class samples while maintaining the underlying data distribution(10–12).

SMOTE effectively balances the class distribution in our dataset. Before applying SMOTE, the majority class (fatal accidents) is overrepresented, potentially skewing the model’s predictions. After applying SMOTE, the minority class (nonfatal accidents) is oversampled, resulting in a more balanced distribution. This balanced dataset can lead to improved model performance, especially in terms of sensitivity and specificity, as the model is now better equipped to learn from both classes.

By employing SMOTE, we aim to mitigate the negative impact of class imbalance and create a more robust and equitable model for our research. This balanced dataset enables our model to learn from both majority and minority classes, leading to more accurate predictions and better insights into the factors contributing to fatal accidents.

**Figure 1:**
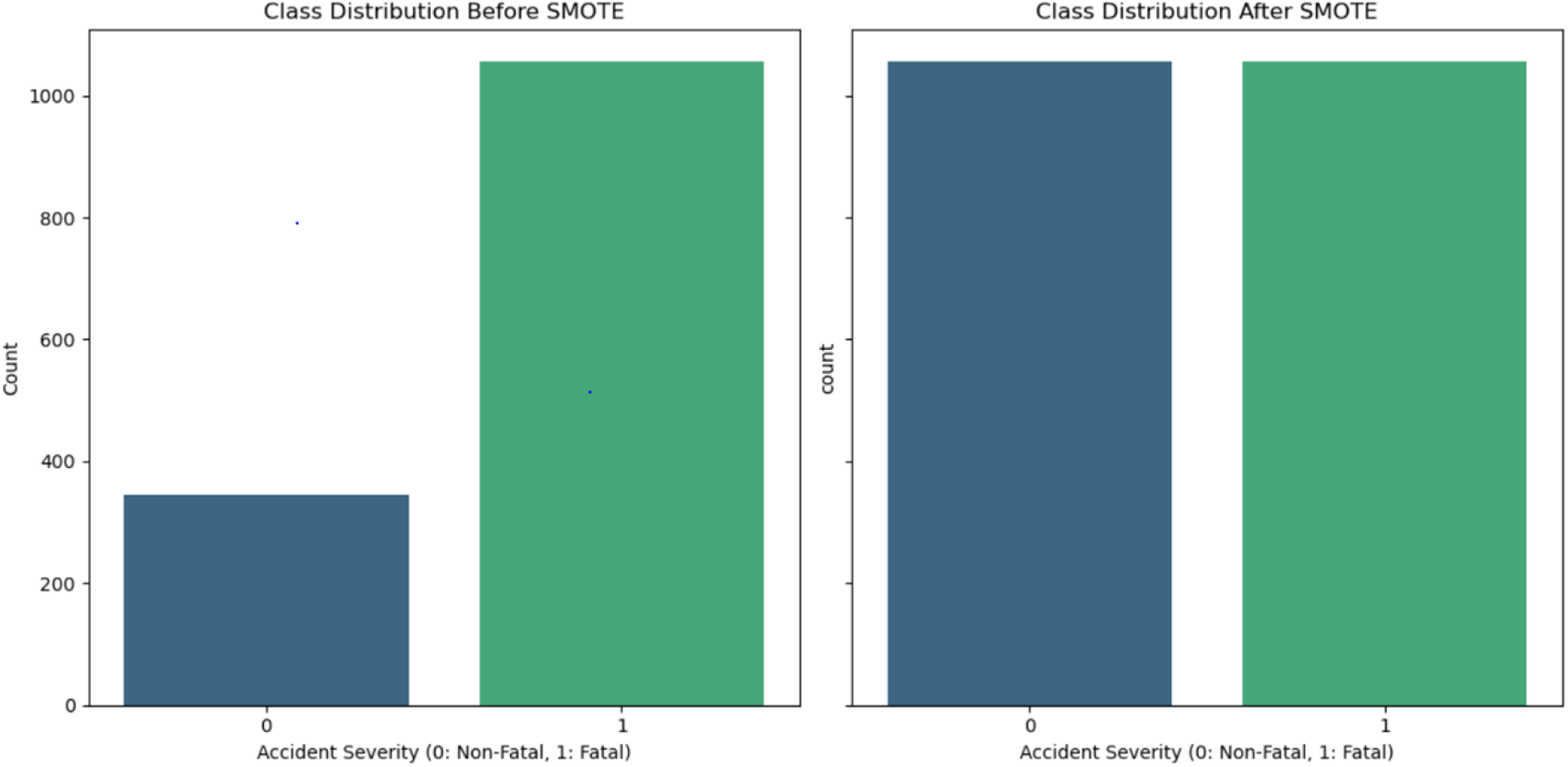
Class distribution before and after SMOTE

### Feature Selection

A correlation matrix was generated to assess the relationships between all the variables in the dataset. Heatmap visualization revealed several notable correlations. Notably, a strong positive correlation was observed between fatalities and injuries, suggesting that accidents involving fatalities are highly likely to also result in injuries. To avoid potential bias and multicollinearity, the “Injuries” variable was excluded from subsequent analyses. Additionally, a moderate positive correlation was found between “Alcohol_Influence” and “Driver_Behavior,” indicating that alcohol-impaired drivers are more likely to exhibit risky driving behaviors. Conversely, a moderate negative correlation was observed between “Seatbelt_Usage” and “Fatalities,” emphasizing the importance of seatbelt use in mitigating accident severity. Other notable correlations included a positive association between “Driver_Fatigue” and “Time_of_Day” (suggesting that fatigue may be more prevalent during certain times of the day) and a negative correlation between “Weather_Condition” and “Traffic_Volume” (indicating reduced traffic in adverse weather conditions). These findings highlight the complex interplay of various factors contributing to car accidents and provide valuable insights for further analysis and targeted interventions.

**Figure 2:**
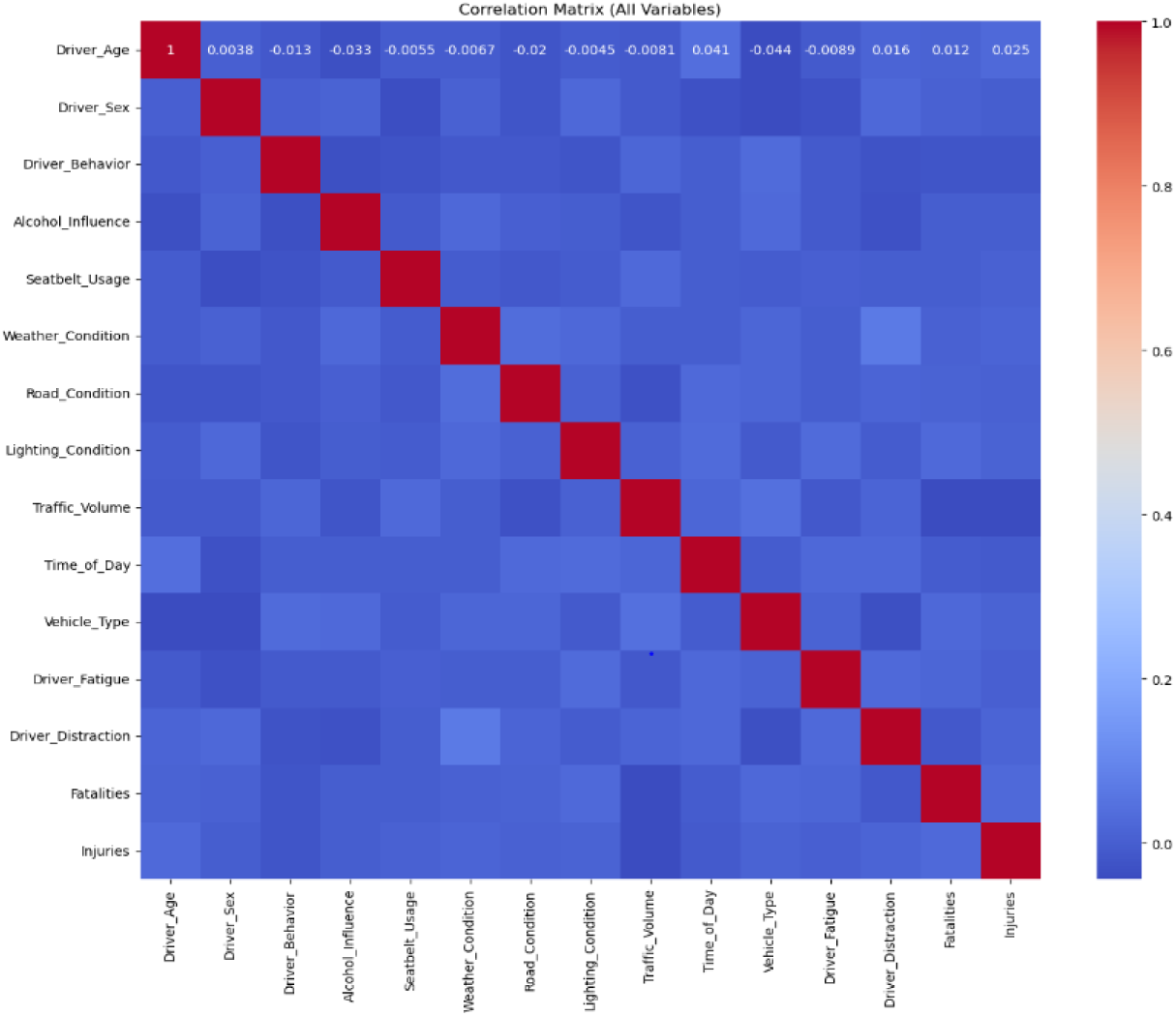
Correlation matrix between variables

To assess the potential for multicollinearity among predictor variables, we calculated the variance inflation factor (VIF) for each variable. The VIF results indicated that all predictor variables presented VIF values below the commonly accepted threshold of 10, suggesting that multicollinearity is not a major concern in this analysis (9). However, the VIF value for “Driver_Age” was slightly greater than those of the majority of the other variables, suggesting a moderate degree of correlation with the other predictors. While this does not necessitate the removal of “Driver_Age” from the model, it warrants careful consideration of its potential impact on model stability and interpretation.

**Figure 3:**
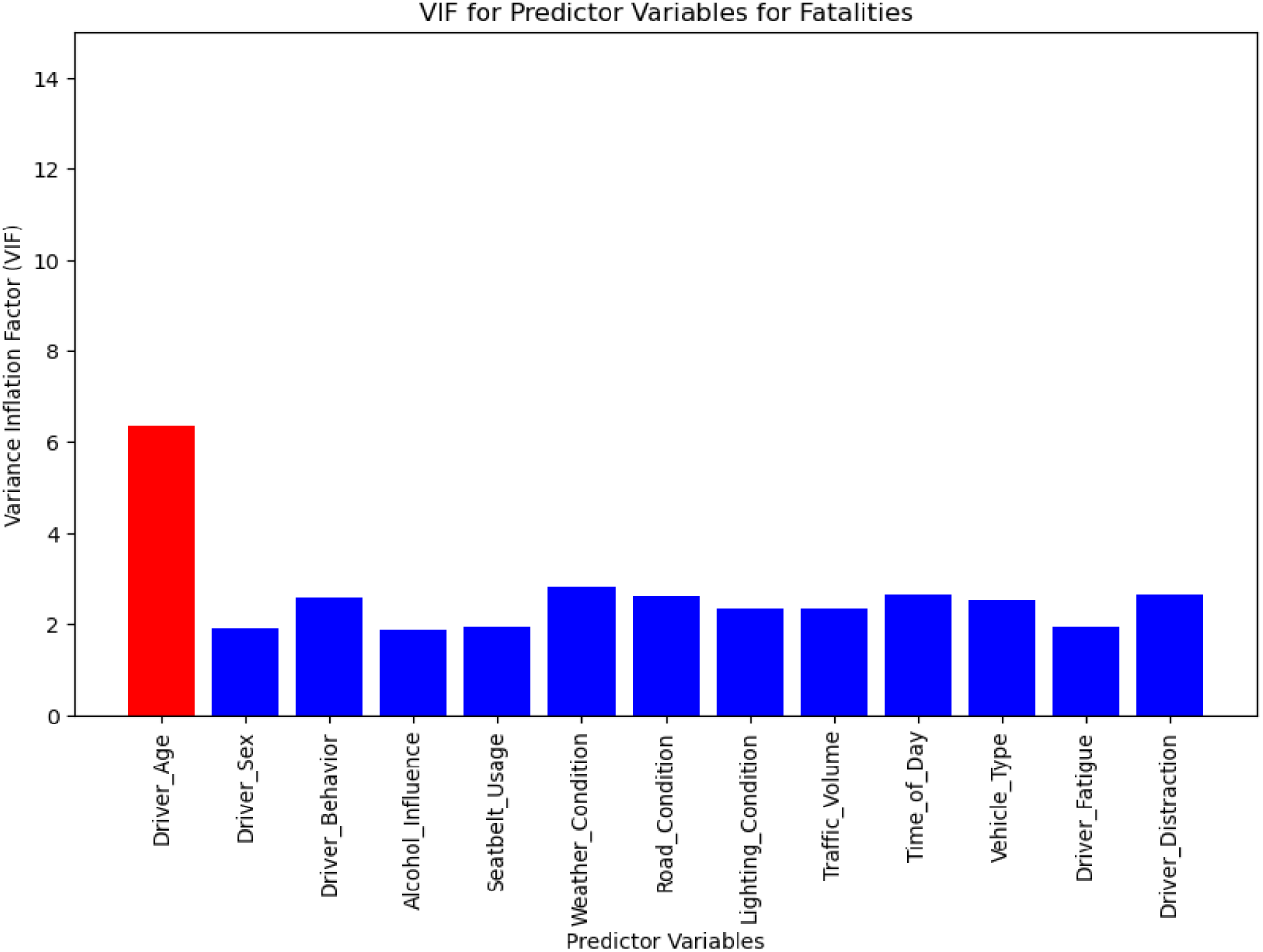
VIFs for predictor variables

### Machine learning approach

For the prediction of accident severity, the study applies ten machine learning algorithms, including logistic regression, decision trees, random forests, gradient boosting, XGBoost, LightGBM, support vector machines, k-nearest neighbors, multilayer perceptron (neural networks), and naive Bayes. These models were selected because of their effectiveness in handling complex, nonlinear relationships and their robustness to high-dimensional data(9,13). The severity of accidents is categorized into two levels: nonfatal and fatal.

Decision trees are popular supervised learning algorithms that create a tree-like model of decisions and their possible consequences. They are known for their interpretability, as the decision-making process can be easily visualized and understood. Decision trees can capture nonlinear relationships between variables, making them suitable for complex datasets(8,14,15).

Random forests build upon the concept of decision trees by creating an ensemble of multiple trees. Each tree in the forest is trained on a different subset of the data and with a random selection of features. The final prediction is made by aggregating the predictions from all the trees in the forest, which typically leads to improved accuracy and robustness compared with a single decision tree(12,13,15).

Gradient boosting is another ensemble learning technique that iteratively builds an ensemble of weak learners (typically decision trees). Each subsequent tree in the ensemble is trained to correct the errors made by the previous trees, resulting in a strong predictive model. Gradient boosting algorithms are known for their high predictive accuracy and ability to capture complex relationships in the data(1,16).

XGBoost (extreme gradient boosting) is an optimized implementation of the gradient boosting algorithm. It incorporates several enhancements, such as parallel processing, regularization techniques, and efficient tree-learning algorithms, resulting in improved performance and scalability. XGBoost is widely used in machine learning competitions and industrial applications because of its high accuracy and efficiency(2).

The LightGBM is another efficient gradient-boosting framework that leverages tree-based learning and parallel processing. It employs techniques such as gradient-based one-sided sampling and exclusive feature bundling to reduce memory usage and improve training speed, making it suitable for large datasets(10,17).

Support vector machines (SVMs) aim to find the optimal hyperplane that best separates data points belonging to different classes. SVMs are particularly effective in high-dimensional spaces and can handle both linear and nonlinear classification tasks(10,11).

K-nearest neighbors (KNN) is a nonparametric algorithm that classifies new data points on the basis of the class labels of their k-nearest neighbors in the training data. The algorithm predicts the class of a new data point by assigning it to the class that is most frequent among its k-nearest neighbors(11,18,19).

A multilayer perceptron (MLP), also known as a neural network, is a powerful model inspired by the human brain. It consists of multiple layers of interconnected nodes (neurons) that process information in a hierarchical manner. MLPs can learn complex nonlinear relationships and are capable of achieving high accuracy on a wide range of tasks(7).

Naive Bayes is a probabilistic classifier based on Bayes’ theorem with the “naive” assumption of independence between features. It calculates the probability of a data point belonging to each class and assigns the class with the highest probability. Naive Bayes is known for its simplicity, efficiency, and ability to handle high-dimensional data(18).

By employing this diverse set of models, we aim to identify the most effective approach for predicting accident severity in Northwest Ethiopia and gain valuable insights into the underlying factors contributing to these events.

### Statistical analysis

Model performance is evaluated via cross-validation techniques to assess the accuracy, precision, recall, and F1 score, and model performance is compared with the AUC‒ROC curve.

Hyperparameter tuning is performed for the outer random forest model to optimize model performance via grid search. The feature importance is also analyzed to identify the most significant factors contributing to accident severity.

All of these studies utilize a machine learning approach to analyze traffic accident data. Data are collected from various sources and undergo thorough preprocessing and exploratory analysis. A range of machine learning models are considered and trained on the data. The performance of the trained models is rigorously evaluated, and the best-performing model is selected for deployment.

**Figure 4:**
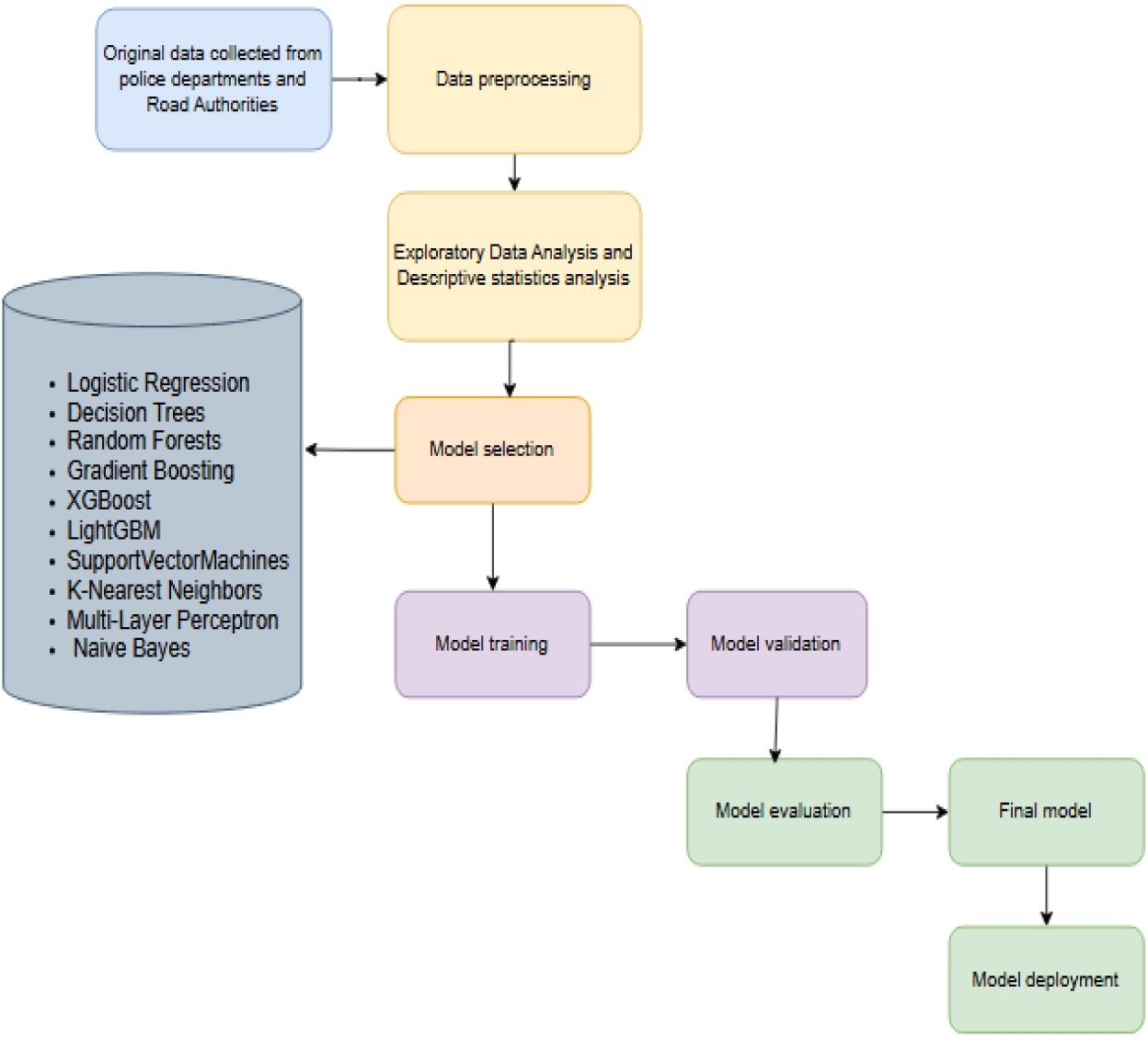
Workflow diagram

## Results

### Descriptive statistics

**Table 1:**
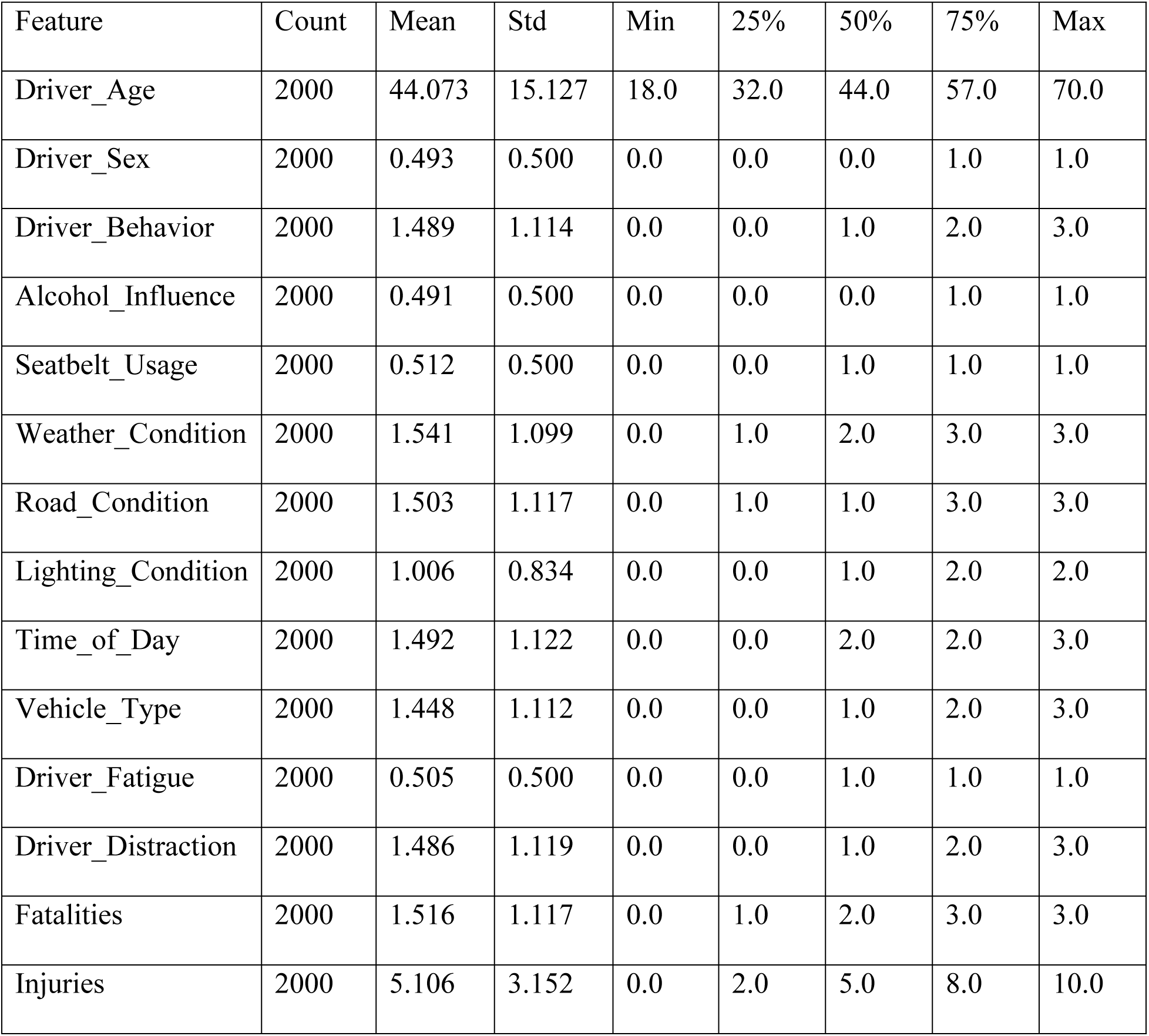
Statistical details of the dataset.

The dataset comprises 2000 car accident records. The average age of the drivers involved in the accidents was 44 years, with a standard deviation of 15.13 years, indicating a considerable age range. Approximately 98.25% of the drivers were male.

In terms of driving behavior, the mean score was 1.49, suggesting a moderate level of risky behavior among drivers. Alcohol influence was observed in 49.1% of the accidents. Seatelt usage was reported in 51.2% of the cases. The mean scores for weather conditions, road conditions, and lighting conditions suggest that a range of environmental factors contribute to accidents. The traffic volume was moderate on average. The majority of accidents occurred during the day (mean Time_of_Day = 1.49). The average number of fatalities per accident was 1.52, and the average number of injuries was 5.11, indicating a significant impact of these accidents.

### Model training results

**Table 2:** Model performance results.

| Model | Accuracy | Precision | Recall | F1 Score | ROC AUC |
| --- | --- | --- | --- | --- | --- |
| Random Forest | 0.784411 | 0.816667 | 0.765625 | 0.790323 | 0.857829 |
| XGBoosst | 0.764511 | 0.776398 | 0.781250 | 0.778816 | 0.854152 |
| LightGBM | 0.771144 | 0.766082 | 0.818750 | 0.791541 | 0.847074 |
| KNN | 0.709784 | 0.846890 | 0.553125 | 0.669187 | 0.819037 |
| Gradient Boosting | 0.759536 | 0.750716 | 0.818750 | 0.783259 | 0.813759 |
| SVM | 0.683250 | 0.727915 | 0.643750 | 0.683250 | 0.732294 |
| Neural Network | 0.673300 | 0.708475 | 0.653125 | 0.679675 | 0.726424 |
| Decision Tree | 0.673300 | 0.718861 | 0.631250 | 0.672213 | 0.676049 |
| Logistic Regression | 0.638474 | 0.678322 | 0.606250 | 0.640264 | 0.673487 |
| Naive Bayes | 0.631841 | 0.668966 | 0.606250 | 0.636066 | 0.672206 |

Model performance was evaluated via a range of metrics, including accuracy, precision, recall, the F1 score, and the area under the receiver operating characteristic (AUC) curve. The ROC curve analysis revealed that the random forest, XGBoost, and LightGBM methods demonstrated superior performance, with their curves closely approaching the top-left corner of the plot, indicating high discriminatory power between severe and nonsevere accidents.

**Figure 5:**
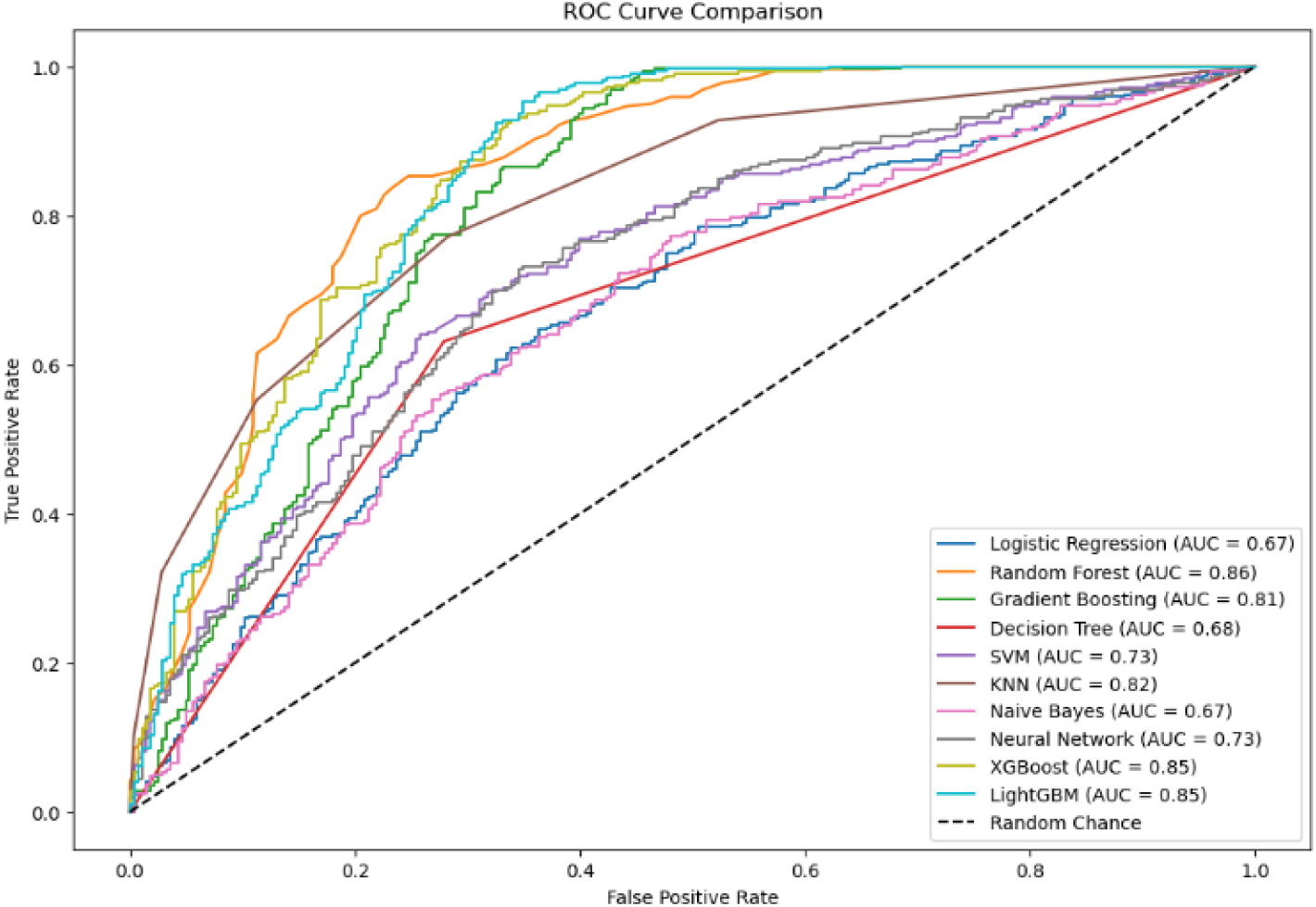
ROC-AUC curve plot for trained models

These findings were corroborated by the model performance metrics. The random forest method exhibited the highest overall accuracy (0.7844), while XGBoost and LightGBM also achieved strong accuracy scores. KNN and random forest demonstrated high precision, whereas LightGBM and gradient boosting exhibited high recall. The random forest and LightGBM achieve a good balance between precision and recall, as reflected by their high F1 scores. These results suggest that ensemble methods, particularly those based on tree-based algorithms, are well suited for predicting car accident severity in this dataset.

**Figure 6:**
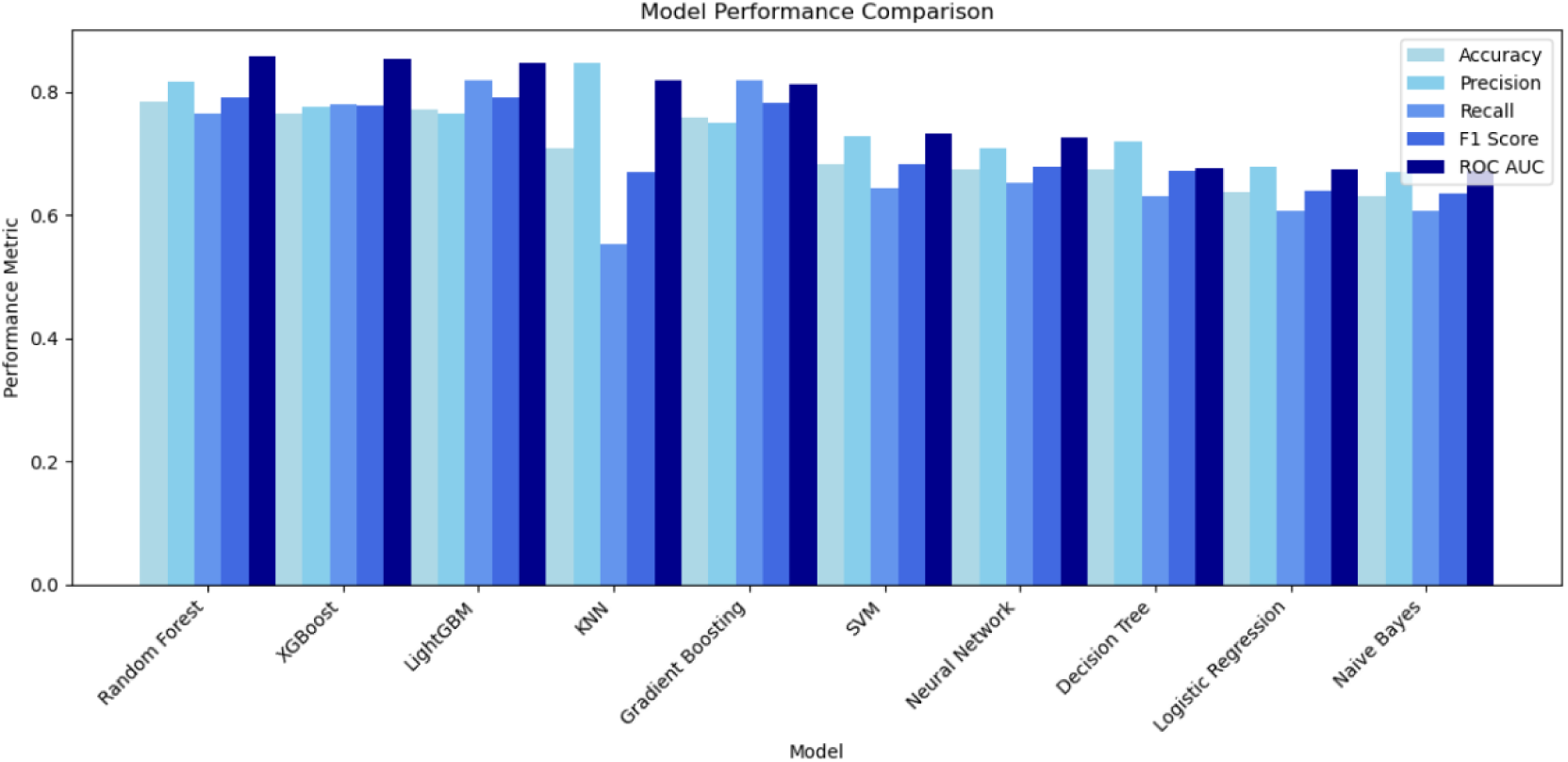
Comparisons of model performance metrics

### Hyperparameter tuning

Hyperparameter tuning was performed via a grid search with 5-fold cross-validation to optimize the random forest model. A range of hyperparameters, including the number of trees, maximum tree depth, minimum samples for splitting and leaf nodes, and the number of features considered at each split, were explored. The optimal hyperparameter combination was selected on the basis of performance on a validation set, resulting in a model with high accuracy, precision, recall, and F1 score, as well as a high AUC score on the ROC curve.

**Figure 7:**
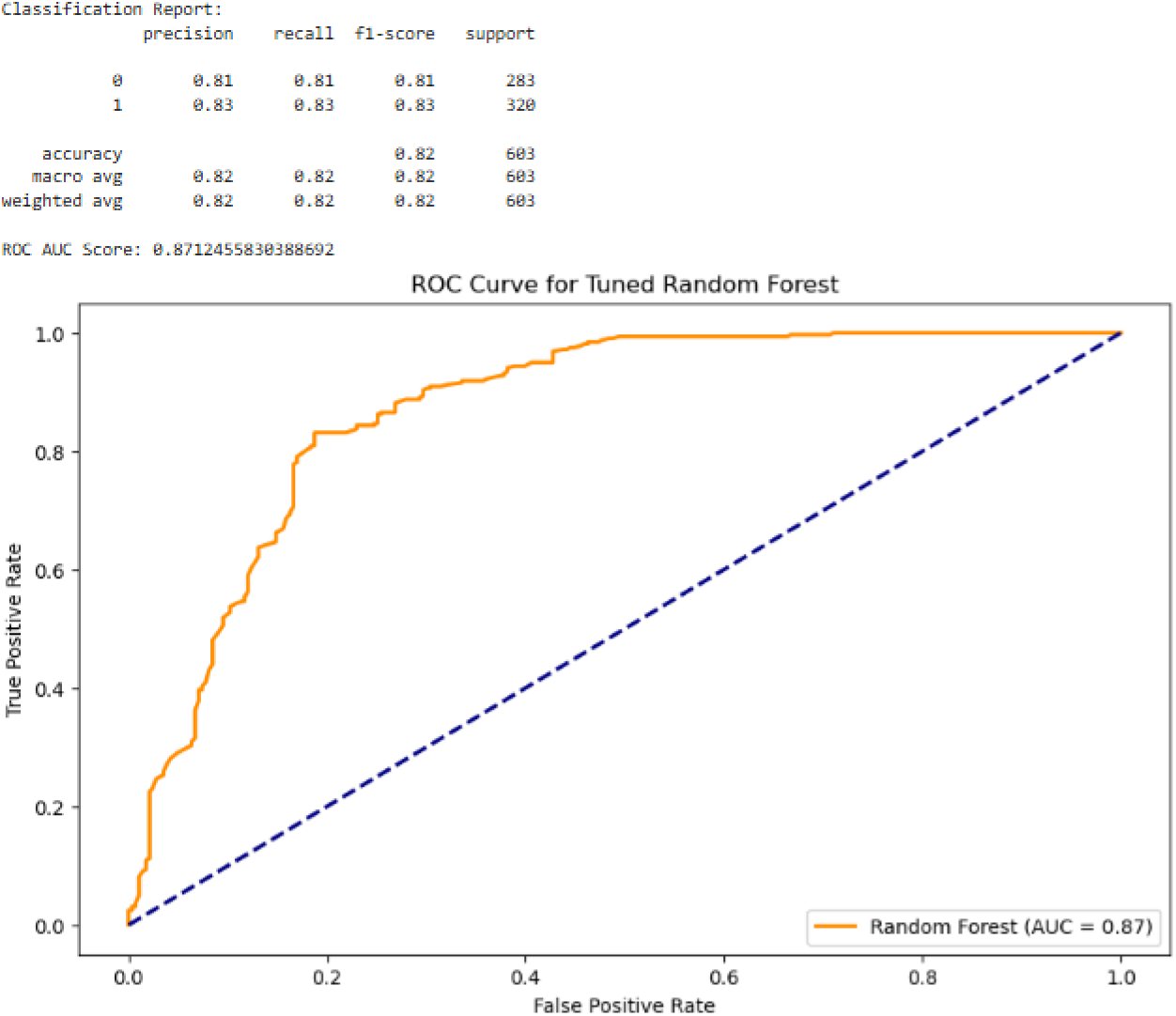
Results of the tuned random forest model

The feature importance analysis was conducted via the random forest classifier. The analysis highlights that Driver Age has the highest predictive power for classifying accident severity, followed by Vehicle Type, Driver Behavior, and Weather Conditions. Variables such as seat belt usage and alcohol influence exhibited relatively lower importance. This ranking underscores the significant role of demographic and situational factors, particularly driver-related characteristics, in determining accident outcomes. These insights contribute to understanding the hierarchical impact of various factors on road accident severity, providing a foundation for targeted interventions and policy recommendations.

**Figure 8:**
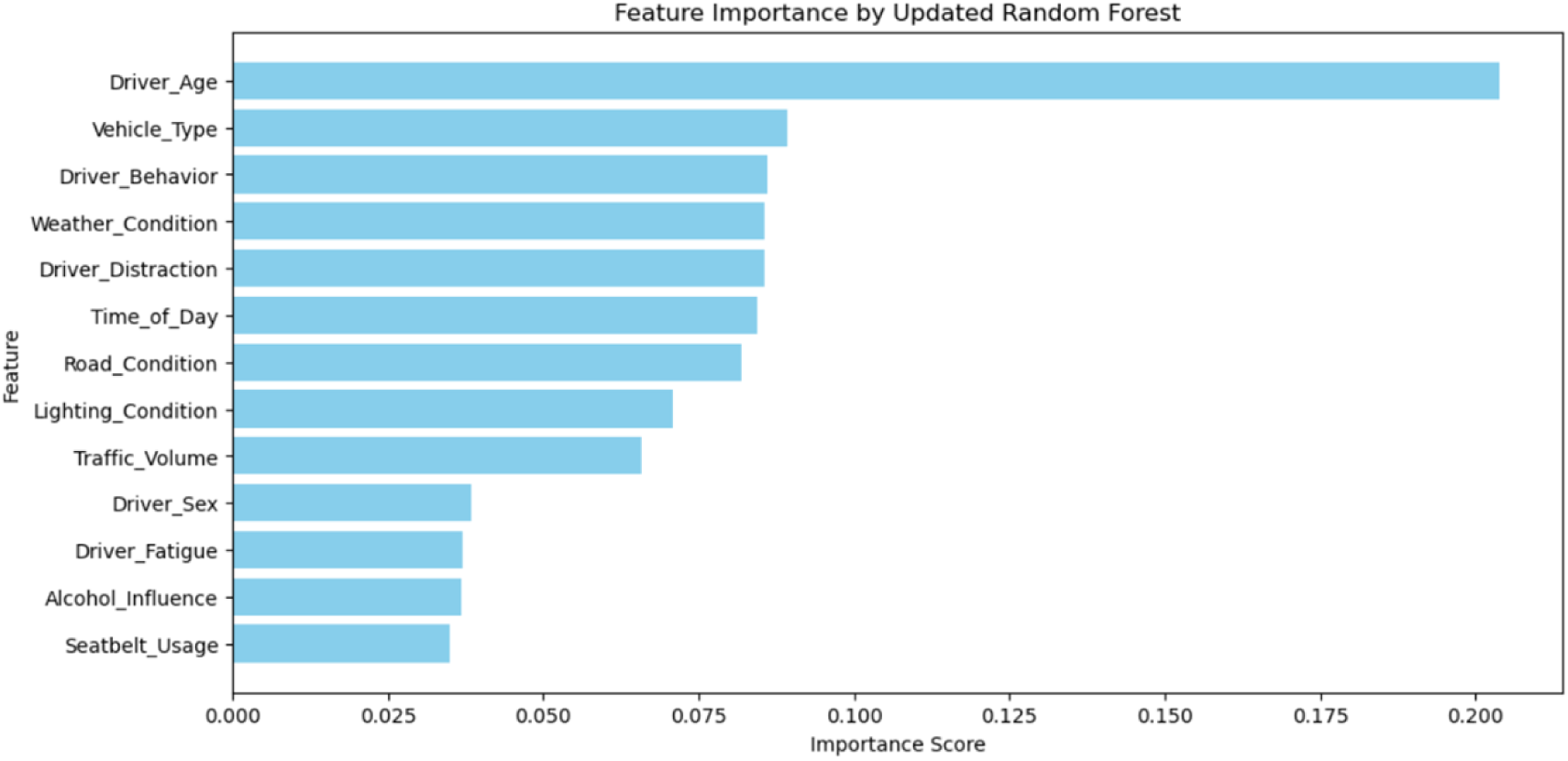
Feature Importance by the Random Forest Method

## Discussion

This study aimed to develop and evaluate machine learning models for predicting car accident severity in Northwest Ethiopia, with a focus on driver, environmental, and road conditions. The results indicated that models such as random forest, extreme gradient boosting, and LightGBM were effective in predicting accident severity, with the random forest model showing the best performance. These findings are consistent with similar studies conducted in other regions, where machine learning models have been successfully used to predict road accident severity on the basis of similar factors(1,11,20).

The findings emphasize the critical influence of driver characteristics, vehicle type, and environmental factors on accident severity. Specifically, Driver Age, Vehicle Type, and Driver Behavior emerged as the most significant predictors. Adverse weather conditions, such as rain and fog, and challenging road conditions, including unpaved or pothole-ridden roads, were associated with a greater likelihood of severe accidents. These results align with studies (5,8,13), which also identified environmental and driver-related factors as pivotal determinants of accident outcomes. The high predictive accuracy achieved by machine learning models, particularly random forests, supports the potential for such models to be applied in real-time accident prediction systems, which could assist in traffic management, emergency response planning, and safety measures(17,21).

Furthermore, the importance of driver age and experience as predictors of accident severity underscores the need for targeted interventions, such as driver education programs and regulatory measures, particularly for younger and less experienced drivers. This finding adds to the body of literature on the role of demographic factors in traffic safety (11,22).

## Limitations of the study

Despite these promising results, several limitations must be considered. First, the study relied on historical accident data, which may not fully capture all relevant variables or changes in traffic conditions over time. The data were also limited to Northwest Ethiopia, which may not be representative of other regions with different road infrastructures or traffic patterns. The sample size, although substantial, may still be insufficient for more granular predictions across diverse subregions.

Finally, while machine learning models perform well, their interpretability remains a challenge. Future work should focus on improving the interpretability and accuracy of these models to ensure that they can be effectively used by traffic authorities and policymakers for preventive measures.

## Conclusions

This study demonstrates the significant potential of machine learning models in predicting car accident severity in Northwest Ethiopia by incorporating key factors such as driver, environmental, and road conditions. The findings indicate that machine learning algorithms, particularly random forests, can effectively predict accident severity, providing valuable insights that could aid in traffic management, emergency response, and safety measures.

The study’s relevance is particularly important for road safety initiatives in Ethiopia, where traffic accidents remain a critical issue. By identifying the most influential factors contributing to severe accidents, such as weather conditions and driver characteristics, this study provides a foundation for targeted interventions, such as improved infrastructure, driver education, and regulatory policies.

Furthermore, the successful application of machine learning models for accident prediction can be a stepping stone toward the development of real-time traffic monitoring systems, enhancing the capacity of authorities to mitigate accidents and improve road safety outcomes.

Overall, this research contributes to the growing body of literature on the application of machine learning in traffic safety and highlights the need for continued exploration in similar contexts to improve predictive accuracy and real-world applicability.

## List of abbreviations

AUC: Area under the curve
F1 score: A statistical measure of a test’s accuracy
GBM: Gradient boosting machine
GPS: Global Positioning System
KNN: K-nearest neighbors
KNN: K-nearest neighbors
LightGBM: Light gradient boosting machine
ML: machine learning
MLP: Multilayer perceptron
RF: random forest
ROC: Receiver operating characteristic
SMOTE: Synthetic minority oversampling technique
Std: Standard deviation
SVM: Support Vector Machine
VIF: Variance Inflation Factor
XGBoost: extreme gradient boosting
XGBoost: extreme gradient boosting

## Ethical considerations

Only the study was conducted using the data that was retrieved. As a result, the data-gathering tool did not contain participants’ names or any other personal information about them. Furthermore, the study was done according to the Helsinki Declaration.

## Availability of Data and Materials

The datasets analyzed during the current study are available from the corresponding author upon reasonable request.

## Authors’ contributions

Abraham Keffale Mengistu conceptualized the study, designed the methodology, collected and preprocessed the data, and performed the analysis. Andualem Enyew Gedefaw analyzed the results and drafted the manuscript.Bayou Tilahun Assaye reviewed and edited for clarity and accuracy. All the authors read and approved the final manuscript.

## Acknowledgments

We would like to thank the East Gojjam traffic police office, Debre Markos city police office, and road safety authorities in Northwest Ethiopia for providing valuable data for this study. We also acknowledge the contributions of the external reviewers for their insightful feedback on the manuscript.

